# Association between social and environmental determinants of health with suicide-related death among veterans

**DOI:** 10.1101/2024.07.02.24309854

**Authors:** Xiange Wang, Wenhuan Tan, Katy (Kaitlyn) Martinez, Benjamin McMahon, Jean C. Beckham, Nathan A. Kimbrel, Silvia Crivelli

**Affiliations:** Applied Mathematics and Computational Research Division, Lawrence Berkeley National Laboratory, Berkeley, CA, USA; Information Systems & Modeling Group, A-1, Los Alamos National Laboratory, Los Alamos, NM, USA; Theoretical Biology and Biophysics, Los Alamos National Laboratory, Los Alamos, NM, USA; Durham Veterans Affairs (VA) Health Care System, Durham, NC, USA; VA Mid-Atlantic Mental Illness Research, Education and Clinical Center, Durham, NC, USA; Department of Psychiatry and Behavioral Sciences, Duke University School of Medicine, Durham, NC, USA; List of people involved in the Million Veteran Program (MVP) Suicide Exemplar Work Group

## Abstract

**Importance:** Social and environmental determinants of health (SDOH and EDOH) may contribute significantly to suicide rates among U.S. veterans.

**Objective:** To identify key predictive variables for assessing suicide-related death rates (SRR), which include suicide deaths, suicide firearm deaths, and suicide non-firearm deaths and vulnerability areas.

**Design, Setting, and Participants:** This case-control study utilized Electronic Health Record (EHR) data, which included demographic and mental health information spanning from January 1, 2006, to December 31, 2016. The base cohort considered all veterans from the VHA outpatient database during the above period. Patients from the base cohort who died by suicide were identified through the National Death Index and considered as cases. Given the significantly larger number of alive patients compared to deceased patients, which caused the dataset to be extremely unbalanced and potentially biased, control participants were selected at a ratio of 4 controls to 1 case from those who were still alive. Cases of suicide-related death were matched with four controls based on birth year, cohort entry date, sex, and follow-up duration. Comprehensive data on social determinants (SDOH), geographic and gun-related factors, quality of access to healthcare, environmental determinants (EDOH), and food insecurity—were gathered from various sources at the midpoint of the study in 2011. Data analysis was carried out from January 2023 to January 2024.

**Exposures:** Suicide-related deaths associated with SDOH and EDOH.

**Main Outcomes and Measures:** A hierarchical clustering method was employed to down-select the large number of variables, while Cox regression models were used to identify key predictive variables for SRR and areas of vulnerability.

**Results:** Out of a total of 9,819,080 veterans, 28,302 were identified as having died by suicide. These cases were matched with 113,208 control participants. The majority of the cohort was male (137,264 [97%]) and White (101,533 [72%]), with a significant portion being Black veterans (18,450 [13.12%]). The average age (SD) was 64.77 (17.56) years. We found that Social Determinants of Health (SDOH) and Environmental Determinants of Health (EDOH) were significantly associated with an increased risk of suicide. By incorporating SDOH and EDOH into the model, the performance (AUC) improved from 0.70 to 0.73.

**Conclusions and Relevance:** In this study, veterans who died by suicide using firearms exhibited distinct characteristics based on SDOH and EDOH, particularly in gun-related variables, compared to those who died by non-firearm methods. Our analysis indicated that veterans living in areas with more social issues, higher temperatures, and higher altitudes are at a higher risk of all-means suicide. Furthermore, regions such as Montana, Wyoming, West Virgina and Arkansas, characterized by higher gun-owernship are predicted to have the highest vulnerability based on veteran suicide firearm rates. Gun ownership and gun laws grades showed as strong predictors rather than rurality.

**Key points:** *Question:* Are social and environmental determinants of health linked to a higher risk of suicide-related death rates (SRR) among US veterans, and can they help pinpoint areas of vulnerability for these individuals preemptively.

*Findings:* Cox models highlighted the significant social and environmental factors that contribute to suicide outcomes. Individuals who died by suicide using firearms have distinct characteristics compared to those who died by non-firearm methods, including higher rates of gun ownership, living in rural areas, greater distances to healthcare facilities. Both groups live in areas that exhibit higher altitudes, a higher percentage of veterans self-reported as white, reduced income and life quality. The model revealed clusters of high-risk firearm suicide individuals in Montana, Wyoming, West Virgina and Arkansas. These areas are characterized by high gun ownership and weaker gun laws. Conversely, clusters of high-risk non-firearm suicides were found in states like California, Washington, and the Eastern coastal area, including New York, where stricter gun control laws and better economic conditions are prevalent. This study shows that gun ownership and gun laws grade, rather than rurality, are the key factors that distinguish between firearm and non-firearm suicides.

*Meaning:* Results of this study suggest that social and environmental determinants of health are associated with higher risk of suicide-related death rates (SRR) among US veterans.

## Introduction

Suicide-related events affect millions of people in the U.S. In 2020, suicide and nonfatal self-harm cost U.S. over $500B in medical costs, work loss costs, value of statistical life, and quality of life costs.^1^ The rate of death by suicide among veterans is 57.3% higher than the rate among civilians after adjusting for age and sex.^2^

We present a model of suicide risk that integrates individual information including clinical and demographics data from the electronic health records (EHR), with geospatial information, including socio-economic (e.g. poverty, gun ownership), environmental (e.g. temperature, heat index), and geographic factors (e.g. rurality, altitude).

Geographic factors such as altitude have received considerable attention within the U.S. as the Mountain West region has a significantly higher rate of death by suicide compared with the rest of the U.S. This pattern is highly consistent across age, sex, race, and population density. Previous work focused on altitude has shown that there is a strong positive association between altitude and suicide.^3–5^ Many other factors have been hypothesized to potentially explain the ZIP code, county and state differences in suicide rates.^4–5^

Substantial literature has shown an association between suicides rates and rurality. McCarthy et al^6^ showed that rural residence was a suicide risk factor among VA patients, even after controlling for mental health accessibility. They also showed that firearm deaths were more common in rural suicides. Rosen et al^7^ showed that increases of more than 20% in suicide rates were seen in 93% of the most rural counties in the 2005-2015 period, in contrast to 79% of suburban (i.e. large fringe metro) counties and 54% of the most urban (i.e., large central metro) counties. Kreise et al^8^ showed that the association between rurality and suicide rates among veterans is significant. Steelesmith et al^9^ showed that suicide rates were higher and increased more rapidly in rural than in large metropolitan counties. However, in 2020, Shiner et al^10^ performed a retrospective cohort study examining differences in the raw and adjusted annual suicide rate among rural and urban VA users between 2003 and 2017 and concluded that the rural–urban suicide disparity appeared to be driven by differences in the racial composition of rural and urban patients, and not by rurality itself.

Altitude and rurality may be a proxy for another variable: percent of gun ownership. Nestadt et al^11^ observed that the suicide rate increases in rural areas are driven by access to firearms. Studdert et al^12^ showed that handgun ownership is associated with a greatly elevated and enduring risk of suicide by firearm.

Other studies have highlighted the influence of the environmental factors on suicide risk. For example, Mullins et al^13^ show a relationship between high temperatures and suicides. Gao et al^14^ observed an association between increasing ambient outdoor temperatures and completed suicide. Cornelius et al^15^ showed that exposure to sunlight, temperature, air pollution, pesticides, and high altitude increases suicide risk.

Despite all the research, risk factors for suicide are still not well understood. Building on Eric Caine’s work^16^, this work pays as much attention to the socioeconomic and environmental context as to the person. It integrates the individual with the context because like Caine, we believe that understanding and preventing suicide can only be achieved through such integration. To that end, we developed a “integrative, hybrid” predictive model of veteran SRR derived from both individual-level and geospatial features. Hierarchical clustering was used to group highly correlated features. We subsetted patients by firearm/non-firearm suicide to assess which features are more strongly associated with each group.

This retrospective study focused on U.S. veterans who are enrolled in Veterans Health Administration (VHA) care and sought to identify predictors for those at risk for SSR. We examined 11 years of individual and geospatial data from 2006 to 2016. We developed a Cox model to predict those veterans at high risk for suicide from 2017 to 2019 and checked the survival probability based on predicted high-risk and low-risk groups and generated a vulnerability map based on the prediction results. The study facilitates the identification of the most predictive factors in advance so that they can be targeted for intervention thus complementing existing clinical services that address the needs of veterans in distress. To the best of our knowledge, no study has integrated such a comprehensive list of individual and geographic variables in a single model.

## Methods

### Datasets

Independent variables: We categorize the variables used in our models as follows: 1) EHR-based demographics(e.g. age, race, gender), 2) EHR-based clinical (e.g. bipolar disorder, PTSD), 3) SDOH (e.g. census demographics, housing infrastructure, education), 4) geographic and gun related (e.g. gun ownership), 5) quality of access (e.g. duration to nearby VA facility), 6) EDOH (e.g. temperature, pm2.5), and 7) food insecurity^17^ (e.g. food shortage). The study cohort is from 2006-2016, and all independent variables except EHR-based were chosen in 2011, since this was midway in the11-yr period that we used. Variables in the dataset have varying spatial resolutions, including county, ZIP code, and state. Since the finest geographic resolution for the EHR dataset is ZIP code, all datasets have been standardized to the ZIP code level.

The dataset features 400 variables derived from a variety of data sources. Figure 1 shows them organized into a four-ring concentric model^18^. The data includes 4 variables from EHR-demographics, 6 from EHR-health, 282 from Social Determinants of Health (SDOH), which forms the largest segment at 70.5%, 10 geographic and gun-related, 30 from Quality of Access which accounts for 7.5%, 58 from Environmental and Other Health Determinants (EDOH) making up 14.5%, and 10 related to food insecurity. The innermost ring categorizes the primary determinants into groups such as SDOH, EDOH, Quality of Access, and Other (which includes EHR, gun-related, and food insecurity factors). The second ring, termed ‘Macro-determinants’, breaks down these principal categories into finer subsections. The third ring, identified as ‘Microdeterminants’, details specific variables such as socioeconomic status within SDOH, environmental factors within EDOH, and elements pertaining to general or VA access quality. The outermost ring identifies the sources for each variable, providing clarity on their origins and the context within which the data was collected (Figure 1).

**Figure 1.**
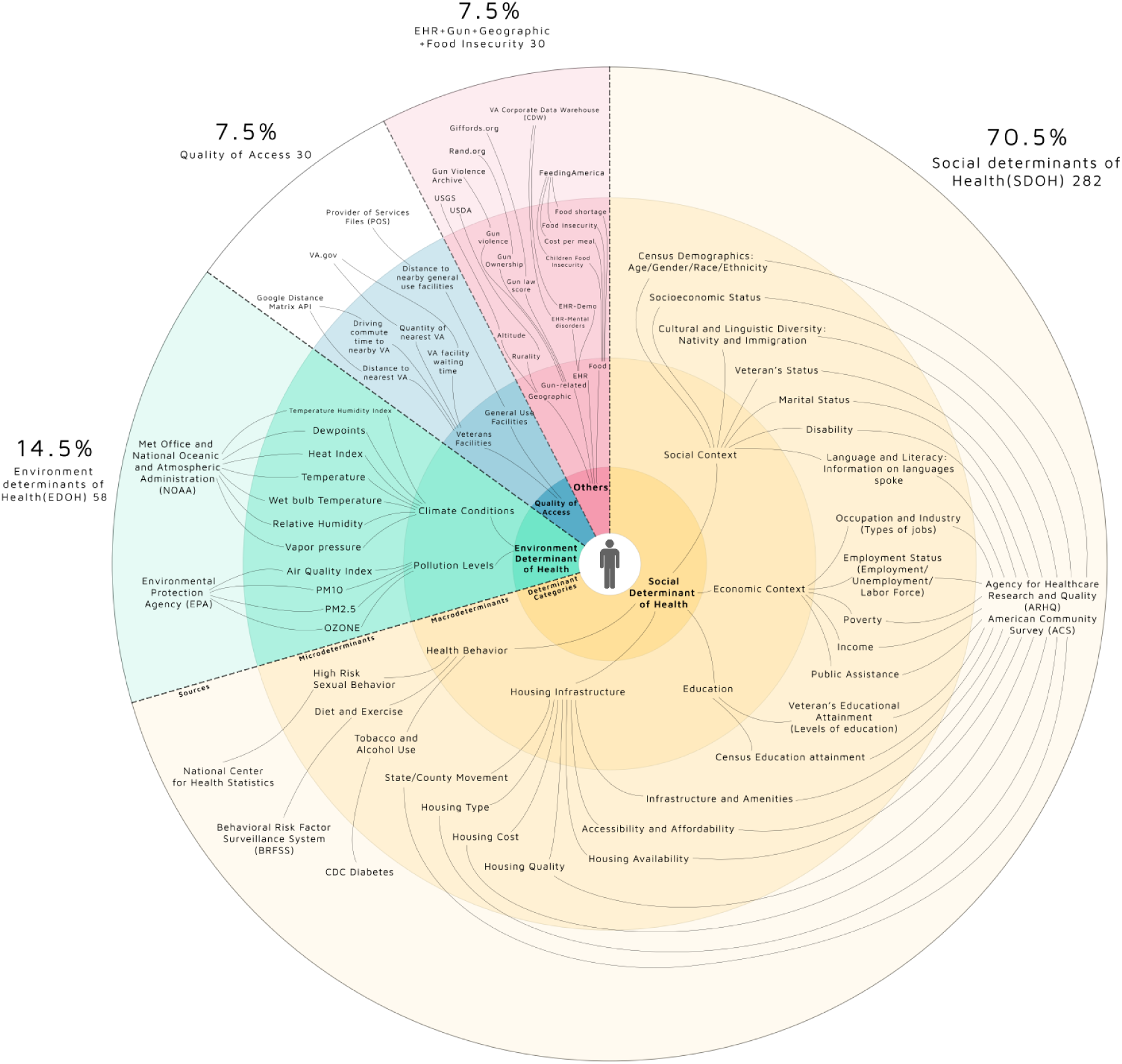
Comprehensive Dataset Summary. Inner Ring: Categorizes main health determinants (SDOH, EDOH, Quality of Access, Other). Second Ring: Subdivides into ‘Macro-determinants’. Third Ring: Details specific ‘Microdeterminants’. Outer Ring: Sources of each variable.

Dependent variables: Using EHR data from 2006-2016, we established annual cohorts of VHA patients. These cohorts consist of patients who received outpatient care or were admitted to a VA facility during this period. The cause of death was based on National Death Index (NDI) search records. To identify suicide deaths, we searched for International Classification of Disease (ICD10) codes X60–X84, Y87.0, and U03*. We then classified these deaths into two categories based on the method used: firearm (codes X72–X74) and non-firearm.

### Statistical analysis

The objective of this analysis was to identify patterns and relationships among individual and geospatial variables using feature reduction algorithms and developed individual models to predict SRR and geographic areas of high risk. The detailed analytical plan involves several steps which are depicted in Supplementary eFigures1 and described in this and next section.

1. Data preparation: data collection, variable selection, data imputation and normalization.
2. Feature reduction to reduce the complexity of the model and the chances of overfitting: selection of a single linkage hierarchical clustering algorithm to produce a dendogram, which is cut at an appropriate level to determine the number of clusters. These results are validated using other methods such as PCA. Selection of clusters’ representatives was based on correlation with the outcome.
3. Developed methods for patient level analysis: Cox proportional hazards model.

### Hierarchical clustering

We utilized a single-linkage hierarchical clustering algorithm (eFigures2 in Supplement 1) that clusters highly-correlated variables. The input includes a set of 400 variables (Figure 1). A multi-criteria optimization method was used to optimize the overall dimension of the representative subset and the information loss due to feature selection. We used the NGSA-II algorithm to get the number of permissible cluster representatives and the correlation threshold which determines the height of the hierarchical tree and partitions the original data into clusters. By using this method, we reduced the original 400 variables to 142 variables which are representatives of the clusters. Some of these clusters consisted of a single variable while others comprised multiple variables that are highly correlated. For the latter, we selected the most representative feature in each cluster by calculating the correlation between the features and suicide rates and selecting the one that is most highly correlated.

### Study population

Due to the large amount of VHA patient data and the fact that suicide is a rare event, we applied a case-control cohort analysis^19^ to our individual analysis. The initial cohort was composed of all veterans for whom the VHA database contains outpatient records between January 1st, 2006 and December 31st, 2016. Each patient’s entry into the cohort was determined as follows: the latest of either the date when the patient had accumulated two years of medical history in the database, the patient’s 18th birthday, or the beginning of 2006. The termination of follow-up was defined as the earliest of the subsequent events: suicide death, death from other causes, conclusion of the last available record for the patient, or the conclusion of the study period (end of 2016). We excluded patients with age over 100 years old.

The cases consisted of all patients in the initial cohort who died by suicide during 2006 to 2016 (n=X). Each case was randomly paired, with replacement, with four control participants from the pool of still-living individuals. The matching criteria encompassed (1) birth year (within ±3 years), (2) year of cohort entry, (3) gender, and (4) duration of follow-up (equal to or longer than that of the corresponding case participant). As per the design, a case participant could serve as a control for another case participant who had died by suicide at an earlier date, and a patient might be a control participant for multiple case participants. However, within each case participant group, the four control participants are not repeated. The index date for each case was defined as the date of suicide, and every control was attributed the same index date as their corresponding case.

### Cox model

We used Cox proportional hazards model to identify important features associated with SRR by examining the coefficient values. The Cox proportional hazards model considers the effect of variables at a time and examines the relationship of the survival distribution to those variables. It is similar to multiple regression analysis, but the difference is that the dependent variable is the hazard function at a given time t (refer to Supplementary eMethods3 for the mathematical description). The Cox model that incorporated the Elastic Net approach was designated as “Cox-EN,” and the model that applied the conventional method for selecting variables was named “Cox.” We utilized both Cox models in our study. We used patient level SSR data which is the time-series data from 2006-2016. For this analysis, we assigned risk time to each VHA user starting on January 1st of 2006 and going through December 31st of 2016; if death occurred prior to the end of the cohort year, risk time was stopped at the date of death. We used the set of reduced variables which resulted from the hierarchical clustering described above.

## Results

Our base cohort consisted of more than 9.8 million Veterans from all VA health care facilities in the United States and its territories; the majority are white, male and 50 years of age or older, with 28,302 deaths by suicide. **Table 1** reports detailed characteristics of the case/control cohort. This cohort consisted of 28,302 case participants and 113,208 matched control participants. Just like the base cohort, most patients in the case-control cohort are above 50 years old, male, and white. We adjusted the Cox model for covariates including race, age, gender, and ethnicity.

**Table 1.**
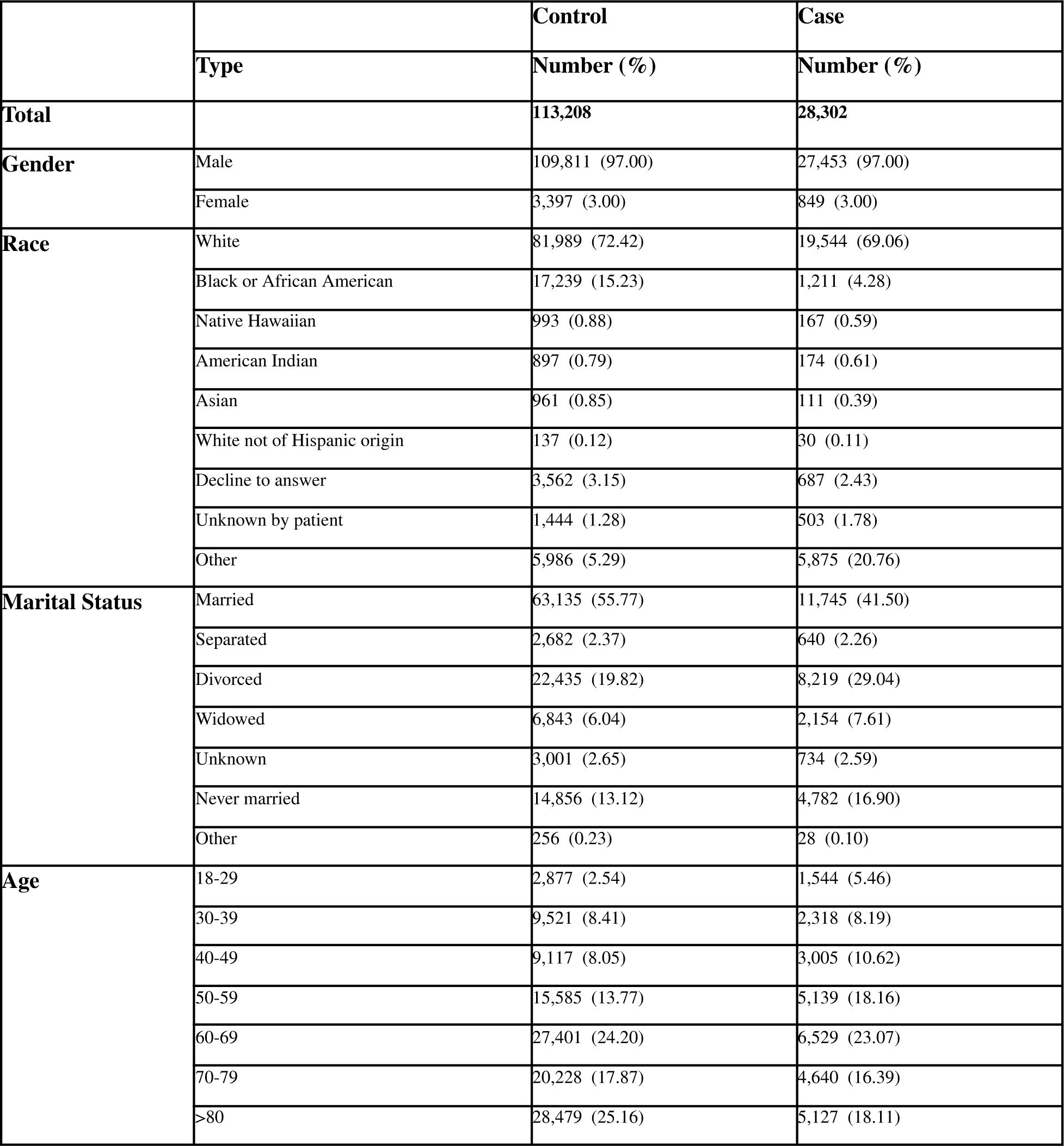
Characteristics of the case-control study cohorts (2006-2016)

We ran a Cox-EN model using the reduced feature set to predict suicide rates. Because the most significant factors predicting death by firearm and non-firearm may be different, we developed separate models to analyze both outcomes.

Table 2 shows that some features are consistently predictive or protective in the 3 groups studied: firearm, non-firearm, and combined. For example, consistently predictive features are food insecurity rate, EHR patient-reported white race, percent of female population divorced or separated, bipolar disorder and altitude. A consistently protective feature is young age (younger than 34).

**Table 2.** Consistently predictive and protective features from suicide, suicide-firearm and suicide nonfirearm using COX-EN.

| **Most Consistent Risk Factors** | **Feature Category** | **Suicide coefficient** | **Suicide Firearm Coefficient** | **Suicide NonFirearm Coefficient** |
| --- | --- | --- | --- | --- |
| Food Insecurity Rate | Food insecurity | 0.8500 | 0.4547 | 0.3864 |
| EHR Patient reporting white race | EHR-Demo | 0.8424 | 0.7426 | 0.8841 |
| Pct of employed working in other services, except public administration | SDOH-Economic | 0.8048 | 0.5361 | 0.5526 |
| Pct of employed working in construction | SDOH-Economic | 0.6052 | 0.6213 | 0.0409 |
| Pct of households with only one occupant | SDOH-Housing | 0.5470 | 0.1678 | 0.0572 |
| EHR Bipolar Disorder | EHR-Health | 0.4695 | 0.2564 | 0.7286 |
| Pct of female population divorced or separated | SDOH-Social | 0.3972 | 0.1943 | 0.4543 |
| Pct of male population married but spouse absent | SDOH-Social | 0.3819 | 0.4288 | 0.0934 |
| Weighted annual food budget shortfall | Food insecurity | 0.3817 | 0.2564 | 0.2541 |
| Pct of employed working in arts, entertainment, recreation and accommodation and food services | SDOH-Economic | 0.3765 | 0.3697 | 0.0232 |
| Pct of population who are private for-profit wage and salary workers | SDOH-Economic | 0.1759 | 0.0563 | 0.1639 |
| Pct of population reporting American Indian and Alaska Native race only | SDOH-Social | 0.1740 | 0.3906 | 0.9017 |
| Altitude | Geographic and gun-related | 0.0980 | 0.0780 | 0.0660 |
| EHR Substance Use Disorder | EHR-Health | 0.0849 | 0.0259 | 0.1434 |
| **Most Consistent Protective Factors** | **Feature Category** | **Suicide coefficient** | **Suicide Firearm Coefficient** | **Suicide NonFirearm Coefficient** |
| Average Heat-Index | EDOH | -0.0677 | -0.0679 | -0.0253 |
| Post-Traumatic Stress Disorder | EHR-Health | -0.1184 | -0.1026 | -0.1066 |
| Pct of children living with a female or male householder | SDOH-Social | -0.3287 | -0.1135 | -0.5461 |
| Pct of children living with a grandparent householder | SDOH-Social | -0.4580 | -0.1212 | -0.7232 |
| EHR Sleep Disorder | EHR-Health | -0.5326 | -0.5022 | -0.5312 |
| Pct of employed working educational service, healthcare and social assistance | SDOH-Economic | -0.5885 | -0.6962 | -0.0403 |
| EHR Patient reporting younger age | EHR-Demo | -0.7622 | -0.7742 | -0.7981 |

Figure 2 shows the most predictive and protective features associated with suicide by firearm. The coefficients are organized in different groups such as demographics (black), SDOH (red), geographic and gun related issues (yellow), economic circumstances (blue), housing infrastructure (green), education (orange), mental health (pink), quality of access (purple), EDOH (cyan), and food insecurity (magenta). For each group, the most significant predictive and protective variables are shown. The height of the bars indicates the magnitude of the coefficient values (predictive above zero and protective below zero).

**Figure 2.**
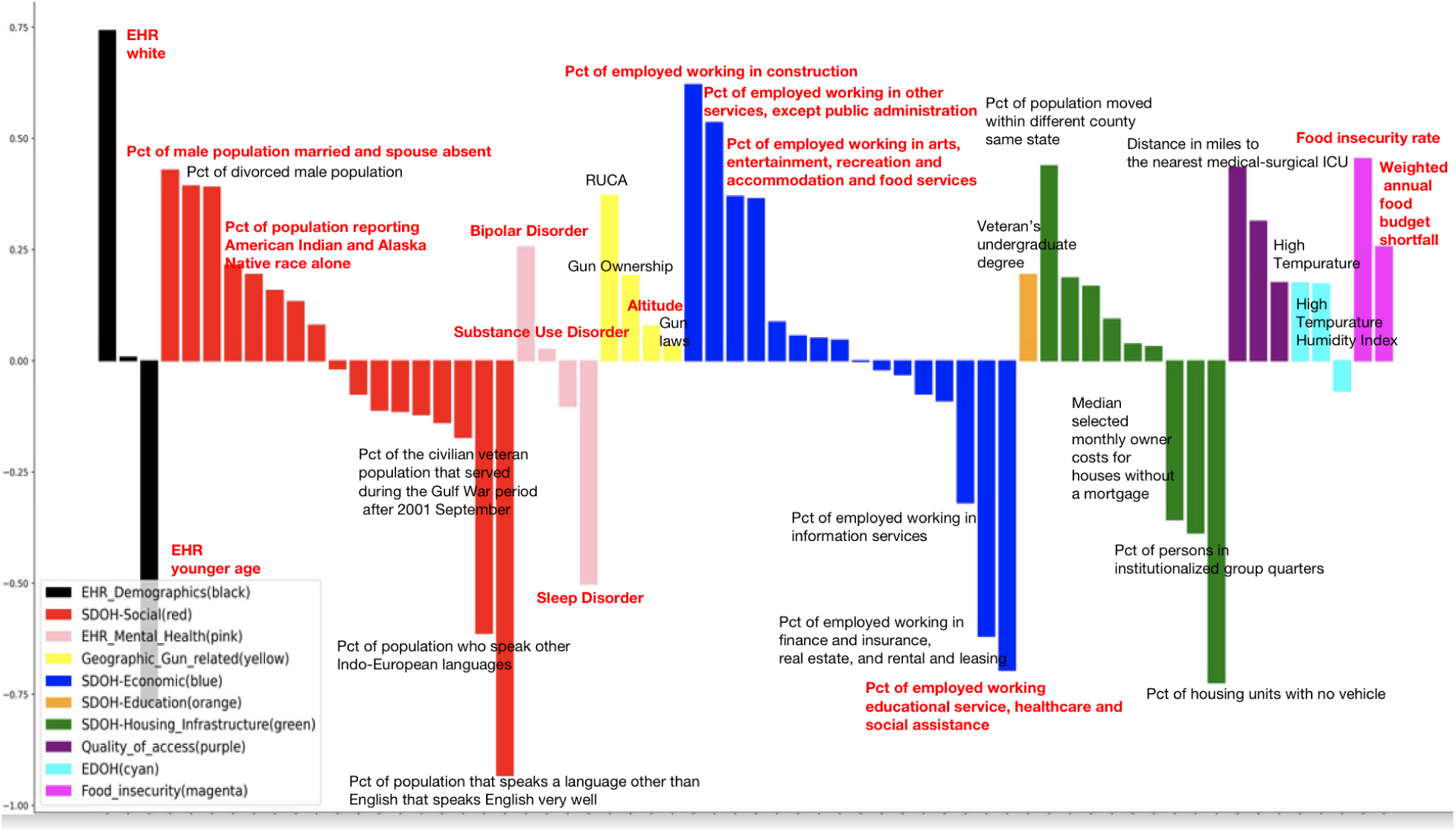
Veteran’s Firearm Suicide COX-EN coefficient by group (2006-16)

Geospatial features associated with poverty, low level of education, distance to facilities with medical-surgical ICU, housing instability (moving within the same state in the past year), rurality, altitude and gun ownership, food insecurity rate, uncomfortable climate changes (higher temperature, temperature humidity index) are predictive of suicide by firearm. Protective features include percentage of people that speak a language other than English and speak English very well, percentage of occupied housing units without mortgage cost, stable jobs).

In the figure, labels marked in bold red indicate factors that are consistently predictive or protective across all suicide-related rates (combined suicide, firearm-related suicide, and non-firearm suicide). Labels in black denote predictors that are unique to firearm-related suicides. For instance, altitude is consistently a predictive factor for all suicide cases, highlighting its significance in the risk of suicide among veterans.

Supplementary eFigures 4 display the most predictive and protective features for suicide, both combined and nonfirearm cases. Supplementary eFigures 5 illustrate the distinctions between firearm and non-firearm suicides. The analysis highlights that gun ownership and the proportion of the population that served during the Vietnam era are predictive factors for firearm suicides but serve as protective factors for non-firearm suicides. Rurality is a significant predictor for firearm suicides but has a zero coefficient for non-firearm suicides. The opposing characteristics of gun ownership between firearm and non-firearm suicides underscore its pivotal role in differentiating the two types of suicides.

The discriminative ability of models was evaluated by the AUC. AUC (Area Under the Curve) measures how well a model can distinguish between different outcomes by comparing the predicted probabilities with the actual binary survival status (alive or dead) of individuals, and by considering the estimated probability of death for censored observations. A higher AUC indicates better model performance. The results demonstrate that the AUC for the COX-EN and COX models using SDOH and EDOH are 0.64 and 0.63, respectively. For the models with EHR mental health features, the AUC values are 0.70. Upon integrating SDOH and EDOH features with the EHR features, the AUC improves significantly to 0.73 and 0.71, respectively. We calculated predicted risk scores for the VHA patients in the study cohort (totalling 140,000), then we sized high-risk and low-risk groups divided according to the risk score based on top 25 percentile and bottom 25 percentile. We observed that the high-risk group has low survival probability compared to low-risk group as shown in supplementary eFigures6.

### Identifying clusters of high vulnerability

We calculated the average individual risk at county level and divided it by the 2017-2019 average VHA county population. This figure was normalized using Min-Max normalization to derive the individual suicide risk values, ranging from 0 to 1, for both firearm and non-firearm suicides. Detailed maps illustrating high-risk counties are presented in supplementary eFigures 7. Utilizing the high-risk values for both firearm and non-firearm suicides, we identified clusters of areas with elevated vulnerability for each type of suicide in Figure 3. Yellow areas indicate high firearm vulnerability, orange areas indicate high non-firearm vulnerability, and red areas indicate high vulnerability for both firearm and non-firearm risks.

**Figure 3.**
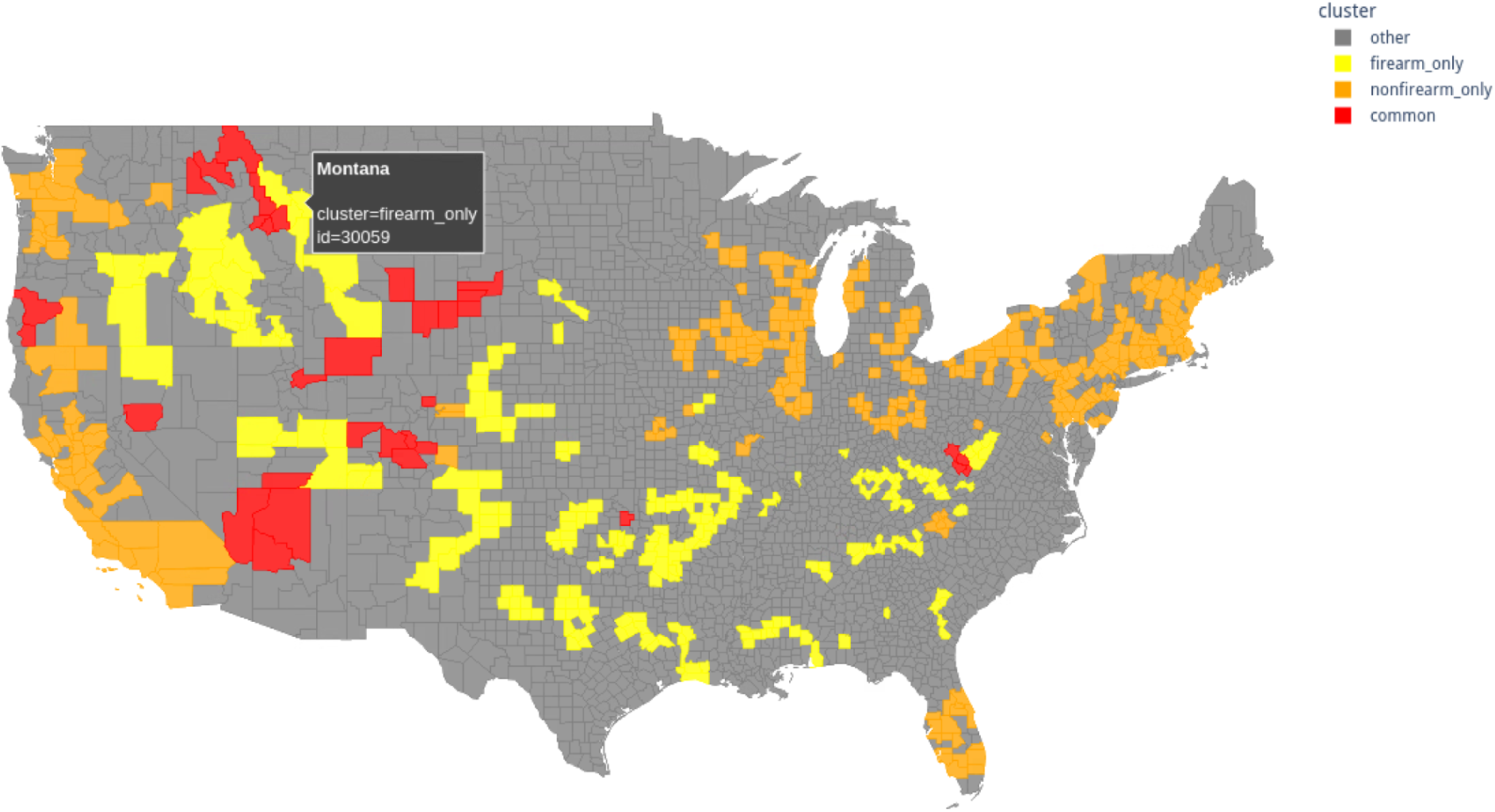
High vulnerability cluster areas for Veterans based on firearm, non-firearm, and both.

In states with high firearm vulnerability, such as Montana, Wyoming, Arkansas, West Virginia and New Mexico, the prevalence of gun ownership significantly exacerbates the risk of firearm-related suicides. These regions not only have the highest rates of gun ownership but also weaker gun-control laws, which may contribute to the increased risk. Conversely, in low vulnerability areas like California and New England, lower gun ownership rates and stricter gun-control laws are observed, which may result in reduced firearm-related suicide risks. Furthermore, areas with lower firearm vulnerability, such as California and New England, tend to face other significant issues, particularly related to drug overdoses, so they have higher non-firearm vulnerability. The stricter gun control measures in these regions correlate with fewer firearm suicides, but the focus shifts to other methods of self-harm, with drug overdose being a prevalent issue. Furthermore, the demographic and socioeconomic factors in these regions may play a role in the prevalence of non-firearm suicides. Urbanization, economic stress, and mental health issues contribute to higher rates of drug overdoses and other non-firearm-related suicides.

## Discussion

This analysis yielded three main outcomes: it identified key social and environmental variables influencing suicide outcomes, examined the characteristics of these variables across different types of suicide, and generated a high-risk suicide score for the years 2017-2019. It integrated Electronic Health Record (EHR) patient-level data with SDOH to identify vulnerable areas for different types of veteran suicides, specifically firearm and nonfirearm. The inclusion of EHR data allowed us to capture detailed patient histories, including diagnoses of mental disorders such as bipolar disorder and substance use disorder, both of which are significantly associated with increased suicide risk rates (SRR). By incorporating SDOH data, we discovered that certain social and environmental factors play a crucial role in distinguishing the vulnerability between firearm and nonfirearm suicides. Gun-related issues emerged as a key factor in differentiating these types of suicides. Veterans with access to firearms or those living in households with firearms are at a heightened risk for firearm suicides. This aligns with previous research indicating that the availability of firearms significantly increases the likelihood of suicide by this method. Furthermore, the study highlights the importance of addressing access to guns in suicide prevention strategies. Policies aimed at reducing firearm access among high-risk individuals, such as veterans with diagnosed mental health disorders, could be pivotal in reducing firearm suicide rates. This approach could involve implementing stricter background checks, promoting safe storage practices, and providing targeted interventions for veterans exhibiting signs of severe mental distress or substance use disorders.

### Limitation

In our suicide analysis, certain EHR mental health disorders, such as PTSD and sleep disorders, consistently exhibit protective characteristics. We have identified individuals with sleep disorders using ICD-9 and ICD-10 codes. To enhance the dataset and improve the accuracy of selecting predictive features, we plan to incorporate Current Procedural Terminology (CPT) codes in future studies. This addition is expected to increase the precision of our data analysis. Furthermore, the predictive or protective features in non-firearm suicides are not as pronounced as those observed in all-means suicides and firearm suicides. To bolster the statistical power for analyzing non-firearm suicides, we plan to include additional years to gather more cases.

Cox models are traditional survival models. In future work we plan to use a random survival model and iterative random forest to detect and account for higher-order interactions and nonlinear relationships among the variables.

We used 2011 SDOH and EDOH features for our survival model, which may not provide the most accurate predictive features due to the lack of temporal effects. To address this, we plan to expand our analysis to include multiple years and use models that incorporate temporal effects. Additionally, we are interested in examining neighborhood and community effects in veteran studies. Therefore, we will utilize geospatial and temporal models, such as hierarchical Bayesian models, to integrate both temporal and spatial effects for more robust results.

## Conclusion

In a previous study we focused on the effect of altitude on suicide deaths, attempts and ideation and showed an association between them. In this study, we showed that veterans who die by suicide with firearms and non-firearms are grappling with other serious traumas and hardships that fuel their attempts to end their lives. We also showed that although altitude is a common predictor for all cases (firearm, non-firearm, and combined), the different groups have different stressors.

Programs aimed to help these veterans usually meet them where they are and respond to their individual needs by improving access to needed resources such as education, employment, housing, and health care. Our study aims to support these programs by developing models that predict high risk at the individual and geospatial levels. We aim to identify not only areas of high vulnerability so that the needed resources can be allocated effectively but also to pinpoint the characteristics of the subgroups and the factors that trigger their attempts.

## Supplemental Online Content

### eMethods1: Details of the data sources

#### EHR-based demographics

We collected individual level demographic data from the EHR. EHR demographic variables include age, gender, race, and ethnicity.

#### EHR-based health data

We collected EHR individual level health data, which include bipolar disorder, anxiety disorders, sleep disorders, substance use disorders, PTSD. The list of International Classification of Diseases (ICD) codes used is as follows:

Bipolar disorder:

ICD-10: F0633, F0634, F30, F31, F340

ICD-9: 2960, 2961, 2964, 2965, 2966, 2967, 29680, 29681, 29689, 30113

Anxiety disorder:

ICD-10: F064, F40, F41, F930, F940, R450, R451, R4582

ICD-9: 29384, 3000, 3002, 30921, 3130, 31321, 31322, 31323, V112

Sleep disorder:

ICD-10: F51, G2581, G470, G471, G472, G473, G474, G475, G4763, G4769, G478, G479, R063

ICD-9: 3074, 327, 33394, 347, 7805, 78604, V694, V695

Substance use disorder:

ICD-10: F10, O9931, F11, F12, F13, F14, F15, F16, F18, F19, F550, F551, F552, F553, G312, F554, F558,F630, Z726, O9932

ICD-9: 291, 2921, 2922, 29281, 29284, 29285, 29289, 2929, 303, 304

PTSD:

ICD-10: F43 ICD-9: 309.81

#### Social Determinants of Health (SDOH)

Our main data source in this category is Agency for Healthcare Research and Quality (AHRQ), which contains a wealth of social and health variables. In addition, we collected social related variables from other sources, including: 1) American Community Survey (ACS) which contains social, economic, housing, and demographic data; 2) County Health Rankings (CHR) and Behavioral Risk Factor Surveillance System (BRFSS) which provides health-related telephone surveys that collect state data about U.S. residents regarding their health-related risk behaviors, chronic health conditions, and use of preventive services.

We divided the SDOH variables into 5 categories:

1. Social context (e.g., age, race/ethnicity, veteran status)
2. Economic context (e.g., income, unemployment rate)
3. Education
4. Housing infrastructure (e.g., housing, transportation)

The spatial levels for this dataset are ZIP code and county level.

#### Geographic and Gun Related

We used the National Altitude Dataset (United States Geological Survey) to get ZIP code altitude data. Rurality was assessed by the rural-urban commuting area (RUCA) codes which classify U.S. census tract using measures of population density, urbanization, and daily commuting at ZIP code level (https://www.ers.usda.gov/data-products/rural-urban-commuting-area-codes/).

We used numerous data points related to firearms such as crime grade (https://crimegrade.org/) from which we obtained the property/violent /overall crime at city level. We obtained the registered gun shop dataset from ATF - Bureau of Alcohol, Tobacco, Firearms, and Explosives which is point-level. In addition, RAND researchers developed annual, state-level estimates of household firearm ownership by combining data from surveys and administrative sources. We assigned laws and policies point values based on their respective strengths or weaknesses. These points are tabulated and the states are ranked and then assigned letter grades (A-F). We also used gun violence data (https://www.gunviolencearchive.org/) for gun participant information, incident information, gun-use information, and gun-violence information at city-level.

#### Quality of Access

This dataset includes factors such as the quantity of nearby VA or general user facilities, distance, driving commute time, and waiting times for each department at annual level. The VA facilities dataset and waiting time are from the EHR dataset whereas distance and driving commute time are obtained from Google’s distance matrix Application Program Interface. General user facilities are from Provider of Services Files (POS) like distance in miles to the nearest hospital with alcohol and drug abuse inpatient care.

#### Environment Determinants of Health (EDOH)

Data in this category are derived from the Met Office Hadley Centre observations (https://www.metoffice.gov.uk/hadobs/hadisd/v330_2022f/index.html) and National Oceanic and Atmospheric Administration (NOAA). Mean/Max/Min temperature, precipitation data are obtained from NOAA, whereas heat index, dew points temperature, humidity index, temperature humidity index and heatwave data are obtained from Met Office Hadley Centre observations. These climate variables are based on each station and corresponding monitors. The spatial-level for this dataset is point-level.

#### Food Insecurity

Map the Meal Gap data from Feeding America organization^17^ provides yearly food insecurity rate, cost per meal and weighted annual food budget shortfall at county-level.

### eFigures1: Analytic plan

This structured and detailed approach aimed to comprehensively analyze the factors influencing veterans suicide risk by integrating various health and environmental determinants using sophisticated clustering and statistical modeling techniques. Data from an Electronic Health Records (EHR) system and additional data on Social Determinants of Health (SDOH) and Environmental Determinants of Health (EDOH) were merged to form a structured dataset spanning from 2006 to 2016. This comprehensive dataset includes EHR-based demographics, health data, geographic and gun-related factors, quality of access, and food insecurity. Feature reduction was conducted using a single-linkage hierarchical clustering algorithm, a multi-criteria optimization method, and the NGSA-II genetic algorithm^20^ to determine the number of permissible cluster representatives and establish a correlation threshold. Subsequently, a detailed patient-level individual analysis was performed, which included a case-control study and the application of the Cox proportional hazards model. The analysis yielded three main outcomes: it identified key social and environmental variables influencing suicide outcomes, examined the characteristics of these variables across different types of suicide, and generated a high-risk suicide score for the years 2017-2019. This structured approach enabled a thorough analysis and prediction of suicide risk based on a broad set of health and social factors.

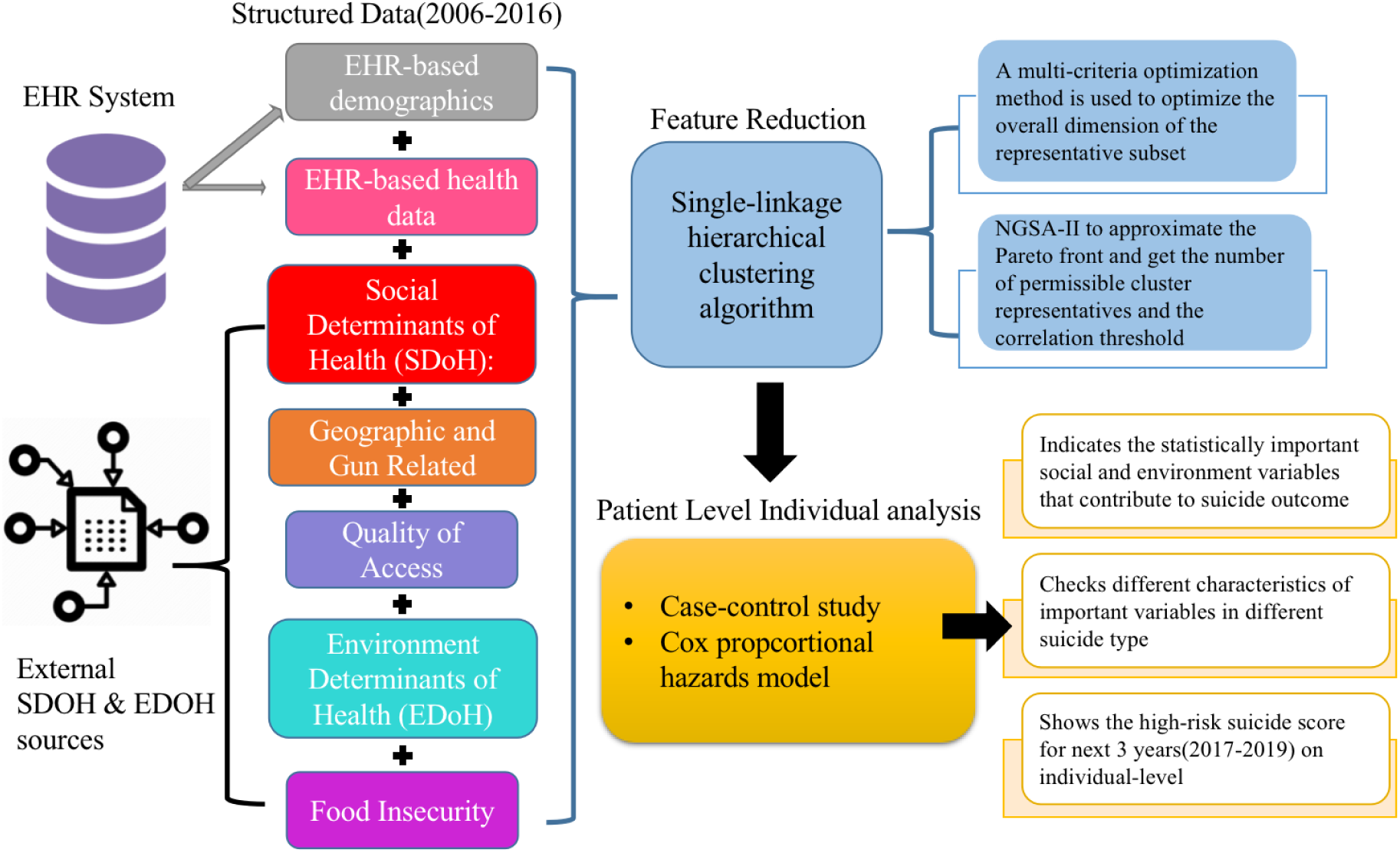

### eMethods2: Features normalization and imputation

#### EHR-based demographics

These variables were categorized as follows:

Age: <18 (0), 18-34 (1), 35-64 (2), >65 (3)

Gender: Female (0), Male (1)

Race: Other races (0), White (1)

Ethnicity: Hispanic (0), Not Hispanic (1)

#### EHR-based health data

For each VHA patient within our cohort (2006-2016), we created separate columns for each disorder (bipolar disorder, anxiety disorders, sleep disorders, substance use disorders, PTSD). We assigned a value of 1 if the patient has an outpatient visit to VHA for the corresponding disorder; otherwise, assigned a value of 0.

##### SDOH

The raw datasets offer both ZIP code and county-level resolutions on an annual basis. For the ZIP code-based SDoH dataset, we applied the min-max normalization method to scale the raw data within the 0-1 range. For the county-level dataset, we utilized the County-ZIP crosswalk file from HUD’s Office of Policy Development and Research (PD&R), which allowed us to correlate county data with ZIP codes before implementing the min-max normalization.

##### Geographic and Gun Related

We correlated the altitude and RUCA values of each patient’s residence ZIP code with their PatientID. We categorized altitude results in our study as follows:

Altitude < 500 meters: 0

Altitude between 500-1000 meters: 1

Altitude between 1000-1500 meters: 2

Altitude > 1500 meters: 3

For RUCA levels, codes 1-3 represent Metropolitan area, 4-6 represent Micropolitan area, 7-9 represent small towns and 10 represent other rural areas. In our study, RUCA values of 1 were labeled as 0, denoting urban areas, while values in the range 2-10 were labeled as 1, signifying rural areas, as it was done in (Steelesmith et al).

We employed various imputation methods for the gun-related dataset. While crime rates and gun violence data are city-specific, we mapped this city data to ZIP codes using the geopy module in Python. All gun shop data was linked to patients based on their ZIP code and the nearest shop. State-level gun ownership and legal score datasets were also correlated with ZIP code data.

#### Quality of Access

We gathered county-level data on distance and driving commute times to nearby VA facilities, utilizing the County-ZIP crosswalk file from HUD’s PD&R to link county data with ZIP codes, followed by applying min-max normalization. Data on general user facilities, sourced from the Provider of Services Files (POS) at the ZIP code level, were also subjected to min-max normalization.

##### EDOH

We allocated daily weather conditions to each patient based on their ZIP code and the proximity of the nearest weather station. Annual figures were determined by averaging the daily metrics and we also calculated the climate bins. For instance, temperature bins were defined as specific ranges (<50°F, 50-60°F, 60-70°F, 70-80°F, >80°F). Each day’s temperature was classified into one of these predefined bins based on its maximum, minimum, or average temperature. The number of days within the year 2011 that fell into each temperature bin was counted. This method was also applied to other climate variables such as the temperature humidity index and heat index.

#### Food Insecurity

For the county-level food insecurity dataset, we utilized the County-ZIP crosswalk file from HUD’s PD&R, associating county data with ZIP codes before applying the min-max normalization.

### eMethods3 Cox Proportional Hazards Model equation

Cox proportional hazards model (Cox model) and Cox elastic net are both used for survival analysis, but they have different approaches to handle variables and regularization.

The Cox Proportional Hazards Model is a semiparametric model used to estimate the hazard ratio for survival data that considers the effect of variables at a time and examines the relationship of the survival distribution to those variables.

Mathematically, the Cox model is written as follows

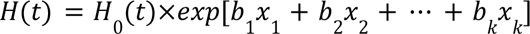

where

- *x*_1_ ··· *x_k_* represent the independent variables,
- *H*_0_(*t*) is the baseline hazard at time t, which is the hazard of an individual having the predictors set to zero.
- By computing the exponential of the regression coefficients *b*_1_ ··· *b_k_*, we can indicate the statistical significance of the relationship between the predictor variable and the hazard rate.

A positive coefficient suggests that an increase in the predictor variable is associated with a higher hazard rate (i.e., increased risk of the event), while a negative coefficient suggests the opposite.

The Cox elastic net model extends the Cox model by incorporating regularization terms to handle high-dimensional data and prevent overfitting. The elastic net regularization is a combination of L1 (lasso) and L2 (ridge) penalties.

The objective function to be minimized in Cox elastic net is:

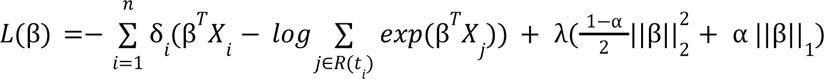

where:

- *L*(β) is the penalized partial likelihood of independent coefficient variable β.
- δ*_i_* is the event indicator (1 if the event occurred for the i-th individual, 0 otherwise).
- *X_i_* is the vector of covariates for the i-th individual.
- *R*(*t_i_*) is the risk set at time *t_i_*.
- λ is the regularization parameter.
- α is the mixing parameter between L1 and L2 regularization (0 ≤ α ≤ 1).

○ When α =1, the penalty is pure L1 (lasso).
○ When α=0, the penalty is pure L2 (ridge).

The elastic net penalty combines the L1 and L2 norms: 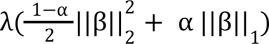

Cox model focuses on estimating the hazard ratio without regularization. It is suitable for low-dimensional data or when overfitting is not a concern. Cox Elastic Net adds regularization to the Cox model to handle high-dimensional data and prevent overfitting. It uses a combination of L1 and L2 penalties, controlled by the parameters λ and α.

### eFigures2: Schematic of the hierarchical clustering algorithm and selection of Pareto solution

We have developed a novel, multi-objective clustering technique aimed at reducing the dimensions of large datasets while preserving the original meaning of the measured variables. This method analyzes the collinearity, covariance, and categorical characteristics of variables to identify the most useful subset of features. Here, utility is defined by achieving a smaller dimension, maintaining variable interpretation, and preserving the intrinsic information of the full dataset. Our approach combines clustering methods and multi-objective optimization to filter and extract attributes, identifying a subset of representative factors for the complete dataset. The correlation-based clustering employed is a unique strategy for partitioning data prior to feature selection or extraction. The utility measures we devise are independent of how the data source is ultimately used, allowing for further variable selection across different applications. The core idea is to leverage the inherent collinearity of the variables, rather than traditional distance measures, to choose the features that best represent the entire set. Specifically, we use an unsupervised hierarchical clustering method to group and classify variables, selecting representative variables to retain within each identified class.

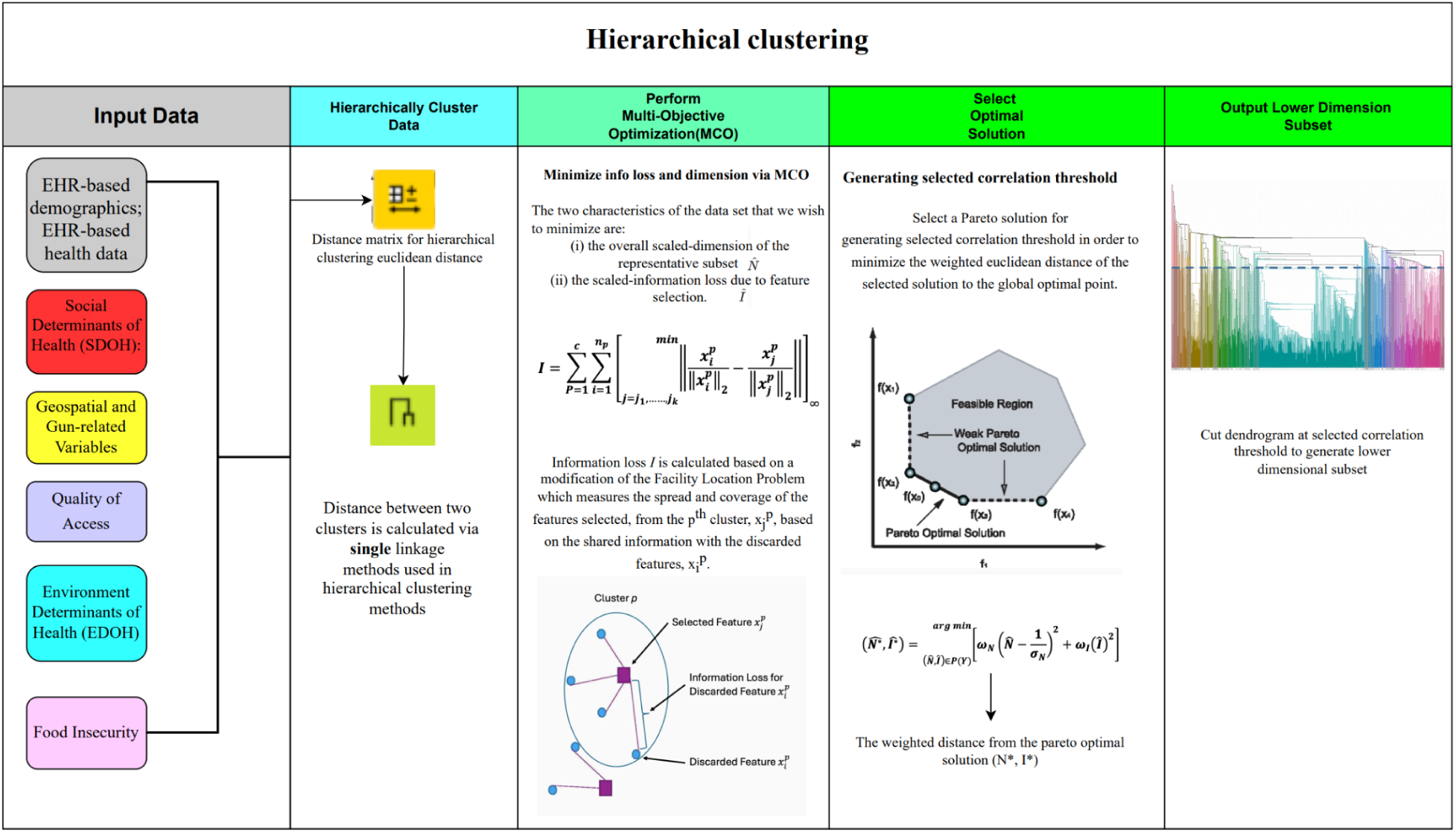

To find the optimal correlation thresholds that correspond to optimal pairs of subset dimension and information loss, we approximated the Pareto front via NSGA-II algorithm. The decision space variable, a continuous range of correlation thresholds, is used to define the multi-objective problem within the algorithm. The number of optimal points N on the pareto front that are estimated impact the convergence of the algorithm, too many and the algorithm will not converge, two few, and the approximation will miss feasible optimal points along the Pareto front. Our current dataset contains 400 features, so we set the N to 160 as nearly half of the size of the dataset. The selection of which optimal point to use out of 160 choices was selected by weighting information loss to dimension of the subset at a ratio of 2:1 by using the green line (see figure below). This green line achieved a correlation value of 0.7336 with 142 features, making it the optimal solution as it maintained a correlation threshold above 0.70 with the smallest feature set among the four points evaluated. The figure below illustrates various weightings: Red (1:2), Blue (1:1), Green (2:1), and Golden (3:1). The red and blue weightings produced correlation values that were too low, while the golden line yielded a subset dimension larger than necessary. This is why we chose the green weighting, as it provided the best balance of correlation and feature count.

#### Information loss vs. representative variable numbers weighting schema

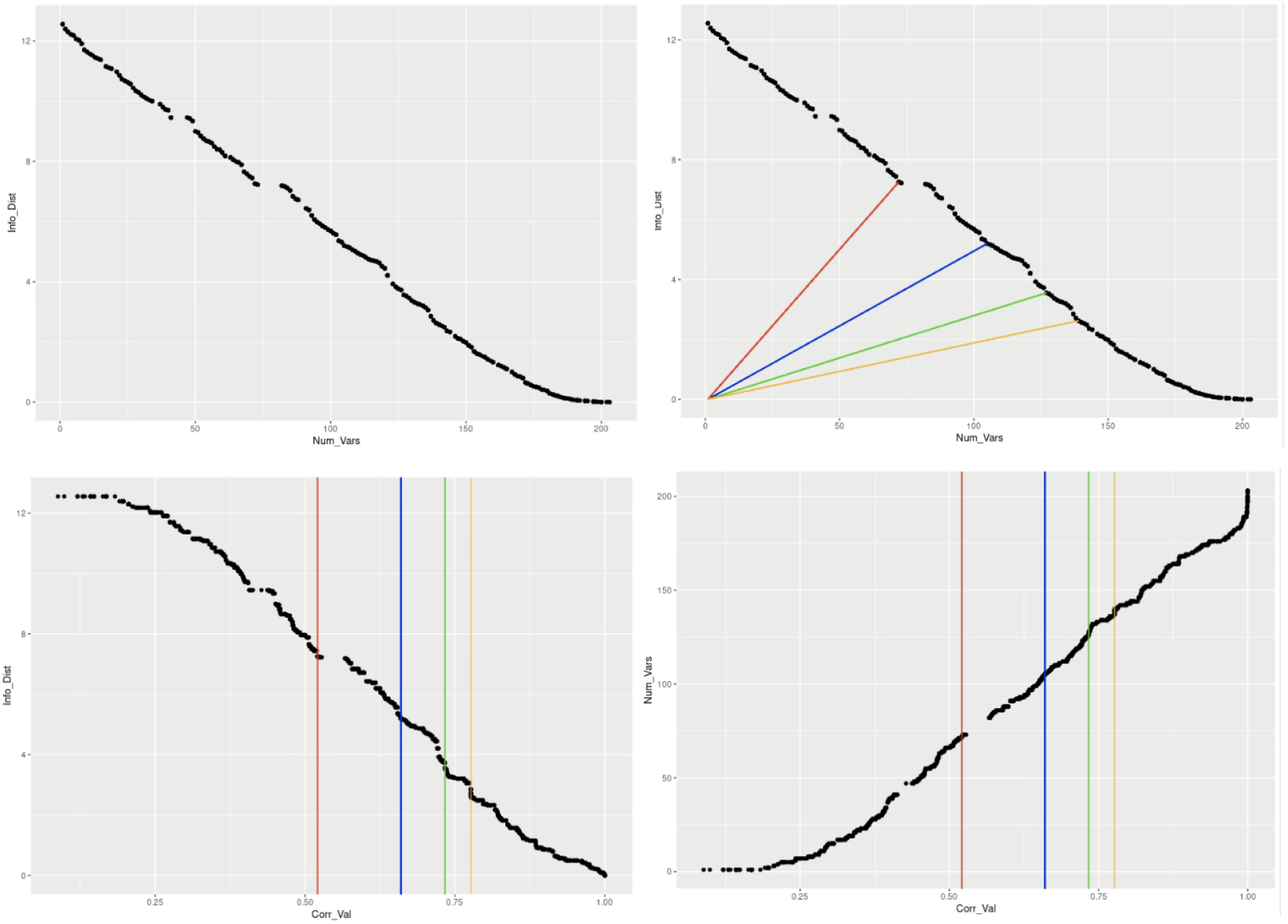

### eFigures3: A schematic of the clustering algorithm and reduced feature group correlation

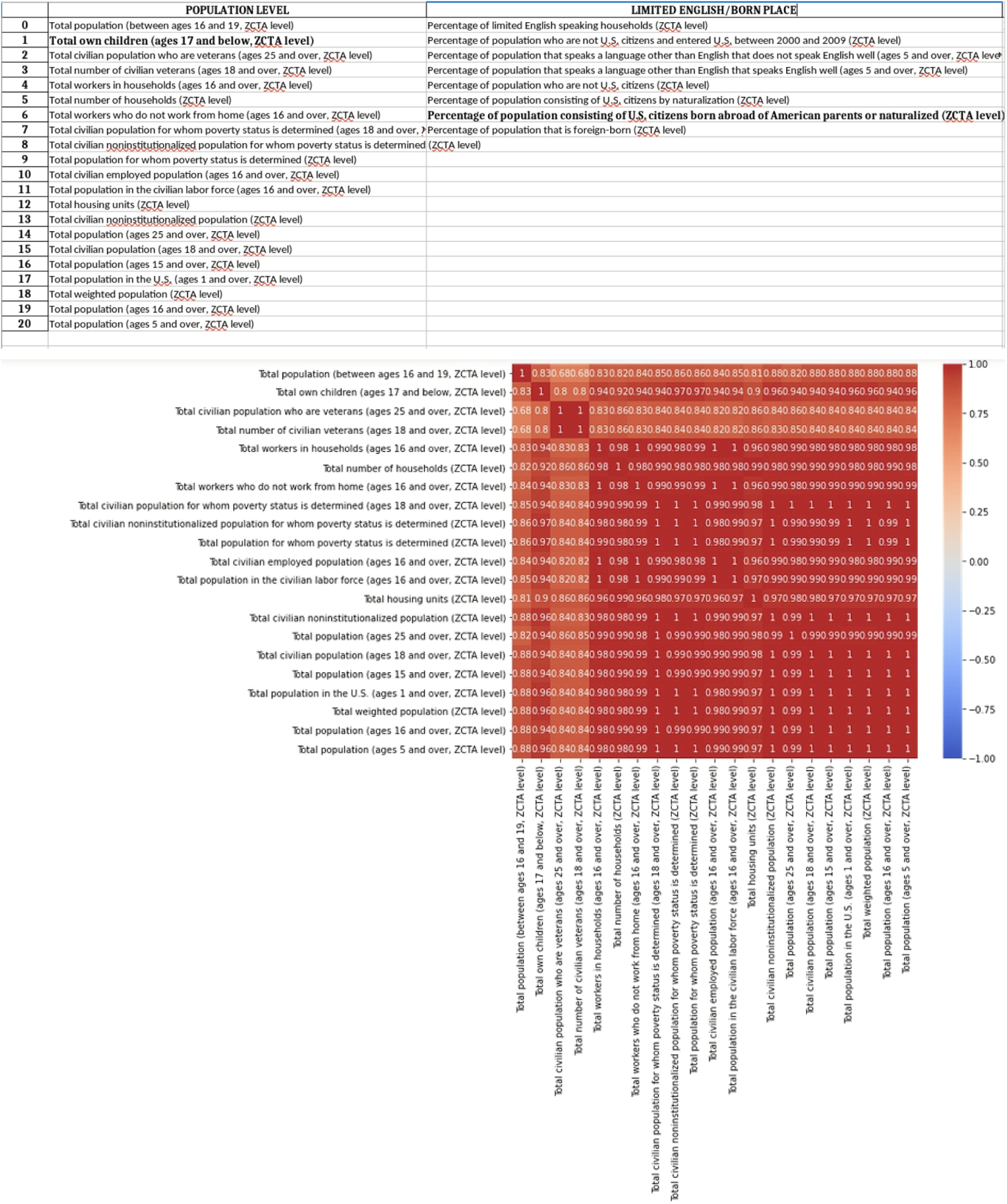

The top panel shows the top 2 clusters that resulted from applying the hierarchical clustering algorithm to the 400 features. They represent topics such as population and citizenship (limited English). The feature in bold text is the most representative feature in each group, selected based on the correlation with suicide rates. The bottom panel represents the heat-map of correlation between each of the variables in the “population” cluster. This heat map shows that all the variables are highly related to each other. This approach helped us remove the redundant features and avoid overfitting when integrating them within the Cox models.

After removing redundant variables, we ensured that the variables within different groups are not highly correlated. This approach helps minimize the risk of overfitting in our model.

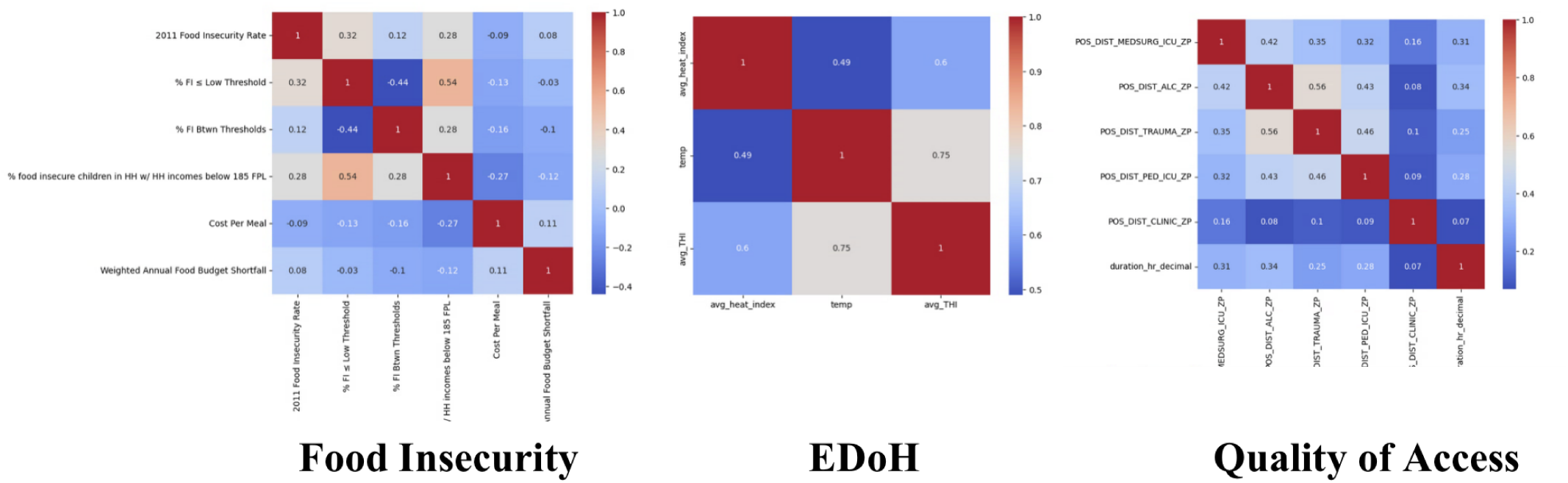

Within the food insecurity group, the variables include:

1. 2011 Food Insecurity (FI) Rate - Indicates the overall rate of food insecurity.
2. % FI ≤ Low Threshold - Represents individuals or households classified within a range that includes low to marginal food security.
3. % FI Between Thresholds - Captures the portion of the population experiencing moderate levels of food insecurity, which is significant but not severe enough to be urgent.
4. % of children in FI HH with HH incomes at or below 185% FPL - Refers to the percentage of children in food insecure households where the household income is at or below 185% of the Federal Poverty Level.
5. Cost per meal and weighted annual food budget shortfall - These metrics provide further insight into the economic aspects of food insecurity.

These five streamlined variables show low inter-variable correlation.

For the Temperature group, the refined features include the average Temperature Humidity Index (avg_THI), average temperature (temp), and average heat index (avg_heat_index), ensuring a comprehensive assessment of climate-related impacts.

In the Quality of Access group, the analysis of correlations among variables related to access to medical facilities shows that all correlations are below 0.6. This group includes:

- Distance in miles to the nearest medical-surgical ICU (POS_DIST_MEDSURG_ICU_ZP)
- Distance to the nearest designated trauma center (POS_DIST_TRAUMA_ZP)
- Distance to the nearest pediatric ICU (POS_DIST_PED_ICU_ZP)
- Distance to the nearest health clinic (POS_DIST_CLINIC_ZP)
- Distance to the nearest hospital with alcohol and drug abuse inpatient care (POS_DIST_ALC_ZP)
- Duration in hours from the centroid of the county to nearby VA facilities (duration_hr_decimal)

### eFigures4: Histogram of predictive and protective features for suicide and suicide by nonfirearm

This plot visualizes a range of predictive features that are categorized by color to represent different groups, each potentially influential in assessing veteran suicide rates. Each bar’s height indicates the magnitude of the feature’s predictive or protective value. The chart is organized into categories such as demographics (black), social factors (red), geographic and gun-related issues (yellow), economic circumstances (blue), housing infrastructure (green), education (orange), mental health (pink), quality of access (purple), environment of determinants of health (cyan) and food insecurity (magenta).

The first plot displays various factors influencing the combined veteran suicide risks (i.e. firearm and nonfirearm). Predictive factors that may increase the risk of veteran suicide are displayed as bars extending above the horizontal axis. These include demographic challenges such as language barriers among non-English speakers and specific age groups. Mental health issues are notably highlighted with the prevalence of bipolar disorder and substance use disorders. Economic factors such as employment in unstable or low-income sectors like arts and recreation are also represented. Additionally, gun ownership appears as a significant geographic factor. Conversely, protective factors that may decrease the risk, represented by bars extending below the horizontal axis, include access to healthcare services and educational attainment. The presence of civilians with higher education degrees and the accessibility of healthcare facilities like trauma centers suggest a buffering effect against suicide risk.

The second plot, which corresponds to veteran nonfirearm suicide risk, also illustrates a variety of predictive and protective factors associated with non-firearm-related suicides, categorized and color-coded by different domains such as demographics, social issues, economic status, housing conditions, and food security. Significant predictive factors include the high prevalence of mental health disorders like bipolar disorder and substance use, demographic vulnerabilities like the divorced female population and certain ethnic minorities, and economic hardships indicated by employment in low-income sectors and high percentages of the population living below the poverty line. Protective factors such as stable jobs, good education, and younger age, lower incidences of the veterans suicide nonfirearm risk.

Overall, these plots effectively displays how complex interactions between various socio-economic, geographic, and demographic factors can influence veteran’s different types of suicide risks, providing essential insights for targeted preventive measures.

#### Veteran’s Suicide COX-EN coefficient by group (2006-16)

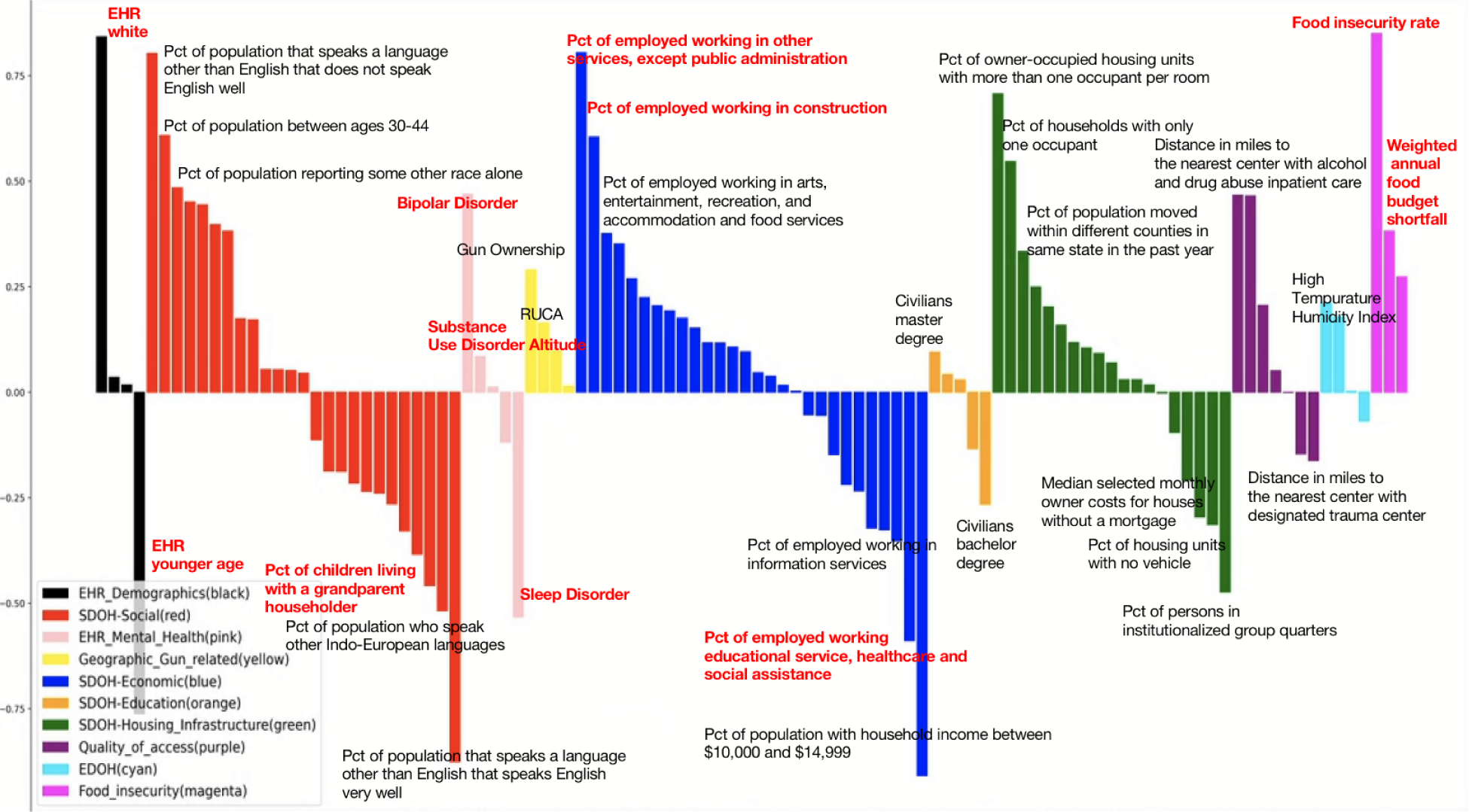

#### Veteran’s Nonfirearm Suicide COX-EN coefficient by group (2006-16)

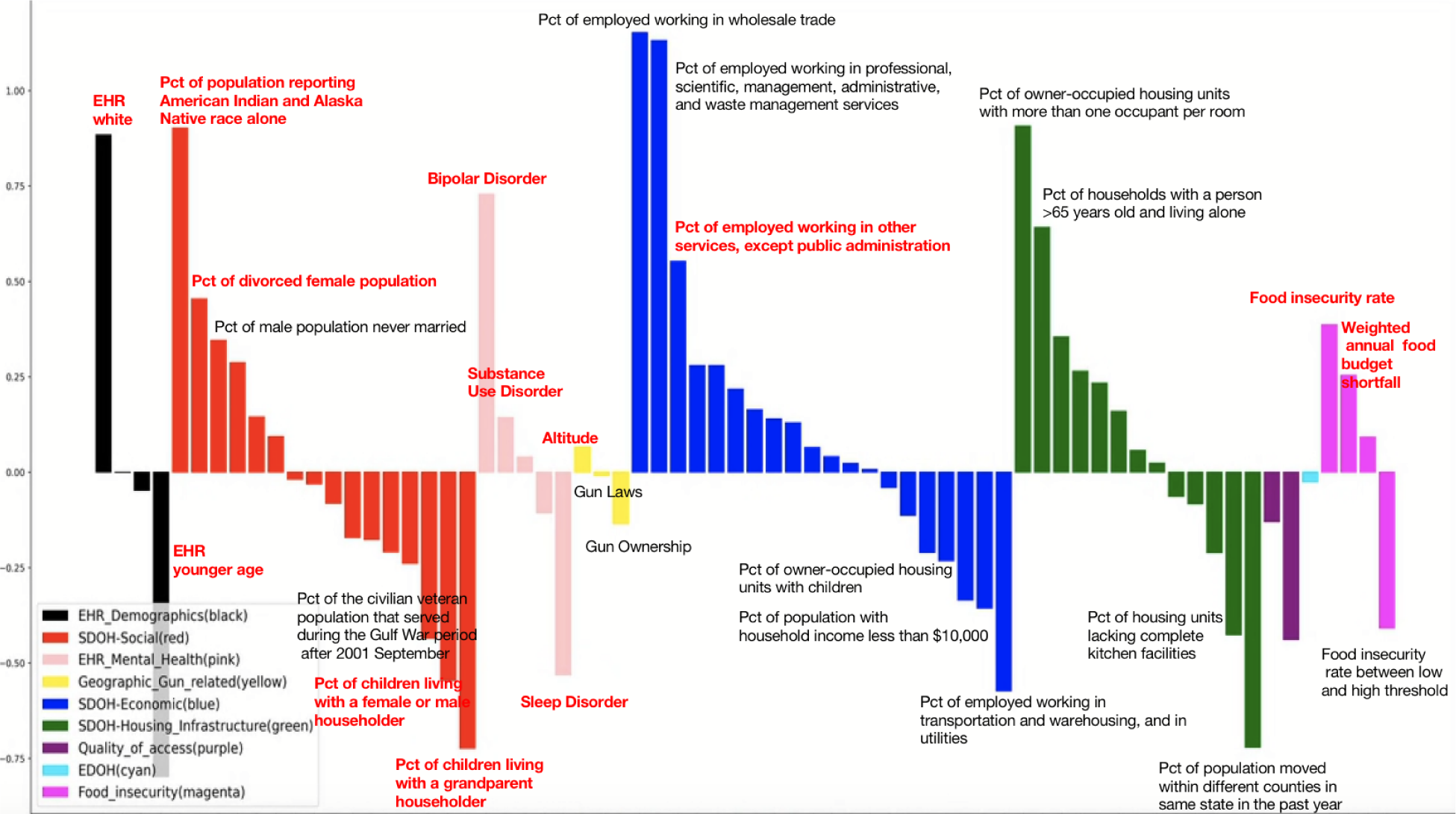

### eFigures5: Difference of predictive and protective features between suicide firearm and nonfirearm

We compare the reduced set of 142 Cox hazard coefficients for the firearm vs. non-firearm suicide deaths models. Variables that are most predictive in both models include Pct of population reporting American Indian and Alaska Native race alone, bipolar disorder, pct of employed working in other services, pct of female population divorced or separated, higher food insecurity rate, weighted annual food budget shortfall. Variables that are most protective in both models include percentage of the civilian veteran population that served during the Gulf War period from September 2001 or later, sleep disorder, EHR patient at younger age. On the other hand, there are variables that have opposite coefficients. For example, Pct of population in labor force are protective for firearm suicide and predictive for non-firearm. Pct of non-hispanic population reporting multiple races, pct of civilian veteran population that served in Vietnam era, pct of population that moved within the same state, are predictive for firearms and protective for non-firearm.

Comparison of coefficients between models for firearm and non-firearm suicide deaths. The x-axis designates the coefficients for each variable in the firearm model, while the non-firearm coefficients are plotted along the y-axis. The color is coded according to the category of variables, as described in the figure legend. The diagonal line is of slope one and can be used to assess the level of agreement in model coefficients. Quadrants are labeled for clarity.

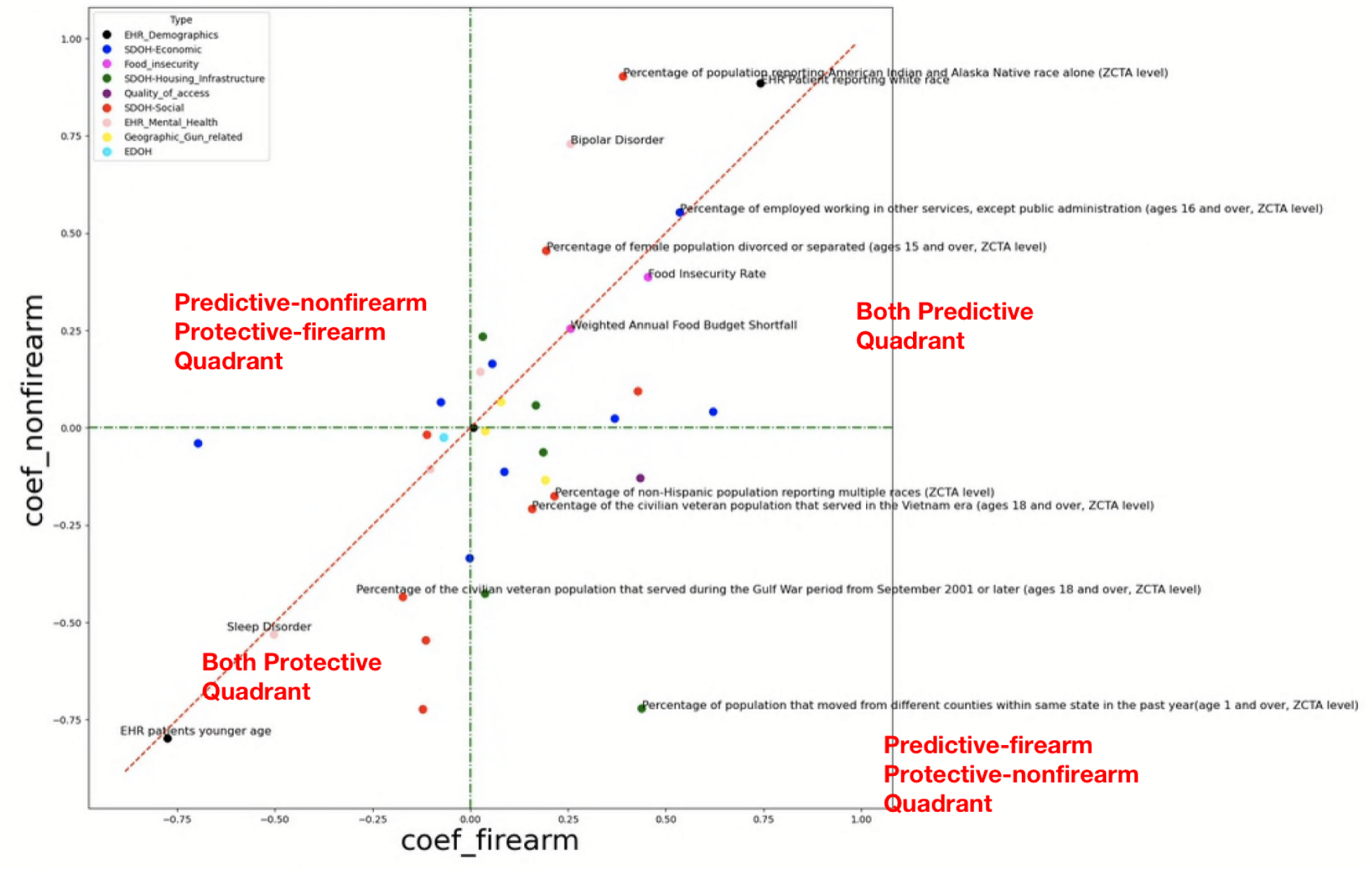

### eFigures6: Survival Probability Curves

The survival curves for high-risk and low-risk groups were estimated using the Kaplan-Meier method, a reliable statistical tool that provides an estimate of survival probability over time. Differences between these curves were statistically assessed using the log-rank test, which is particularly effective for comparing the survival distributions of different groups. The survival curves, categorized according to the calculated risk score, are illustrated below.

Our analysis demonstrates a distinct pattern: individuals in the high-risk group exhibit a significantly lower probability of survival as time progresses. This trend starkly contrasts with that of the low-risk group, whose members show a notably higher survival probability over the same period.

#### Survival Probability Curves

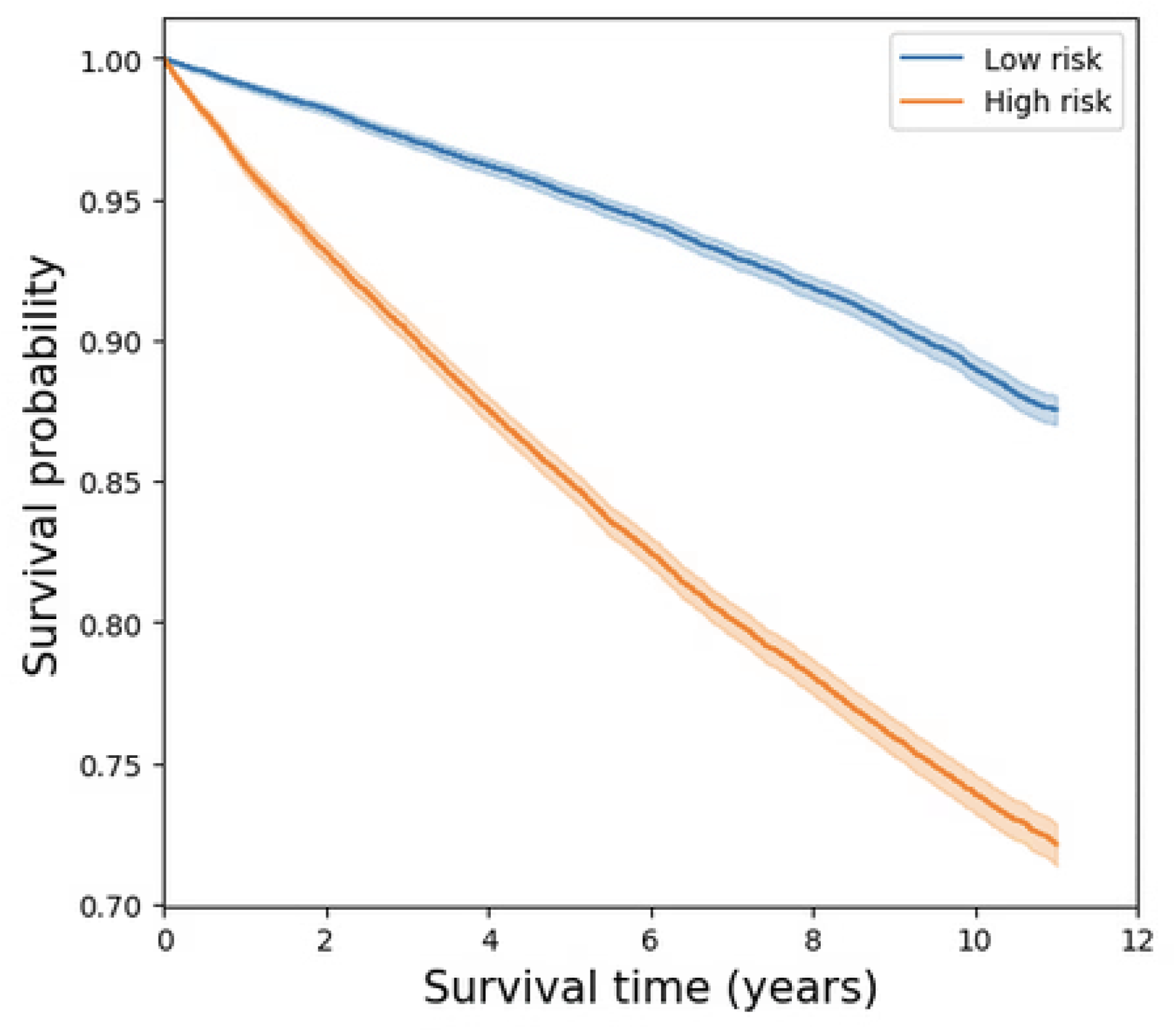

### eFigures7: Vulnerability maps for VHA outpatients suicide, suicide firearm and suicide nonfirearm

As illustrated in the maps, higher VA suicide vulnerability areas are more concentrated in Wyoming, Colorado, Oregon, and Nevada. These regions are predominantly rural and high-altitude, factors that may contribute to increased vulnerability. Additionally, high firearm suicide vulnerability areas are notably concentrated in Montana, Wyoming, Arkansas, West Virginia, and New Mexico. We can see that the areas of Arkansas and West Virgina (circled in black in the firearm map) have higher risk rates compared to the all-means suicide vulnerability map. Those are regions where gun ownership is more prevalent and gun-related issues are more acute. In contrast, higher non-firearm suicide vulnerability areas, notably suffering from issues like drug overdose, are found in states such as Washington, California, New York, and New Jersey, which have been circled in black in the nonfirearm map.

These maps aid in visualizing the potential high suicide risk among veterans and underscore the need to consider diverse factors such as altitude, rurality, and various social, gun-related, and drug-related issues in future studies.

More details from the suicide vulnerability maps reveal that states like Wyoming, Colorado, Oregon, and Nevada face specific social issues that exacerbate the risk. For instance, Wyoming, with one of the highest rates of gun ownership in the U.S., also grapples with social isolation due to its rural nature and limited access to mental health services. Economic challenges, such as job availability and healthcare access, further compound these issues.

Colorado’s higher altitudes are associated with increased rates of depression and suicide. The state struggles with substance abuse and significant economic disparities between urban and rural areas. Oregon, similarly, is challenged by inadequate mental health services, particularly in rural areas, and high rates of substance abuse, including opioid addiction. The substantial homeless population in Oregon correlates with higher rates of mental health issues.

Nevada shows stark differences in access to healthcare and mental health services between urban areas like Las Vegas and its rural counterparts. Economic instability, high unemployment rates, and prevalent substance abuse in urban centers contribute to the elevated suicide risk.

In contrast, areas like California and the New England states, which show lower suicide vulnerability, generally have better access to mental health services and resources. Higher urbanization rates in these areas provide more social connectivity and support networks, while greater economic opportunities and resources help reduce stressors related to financial instability. However, California faces a high number of homeless and substance abuse which increases suicide by non-firearm vulnerability.

#### High Suicide Vulnerability Areas Based on Individual VHA Data

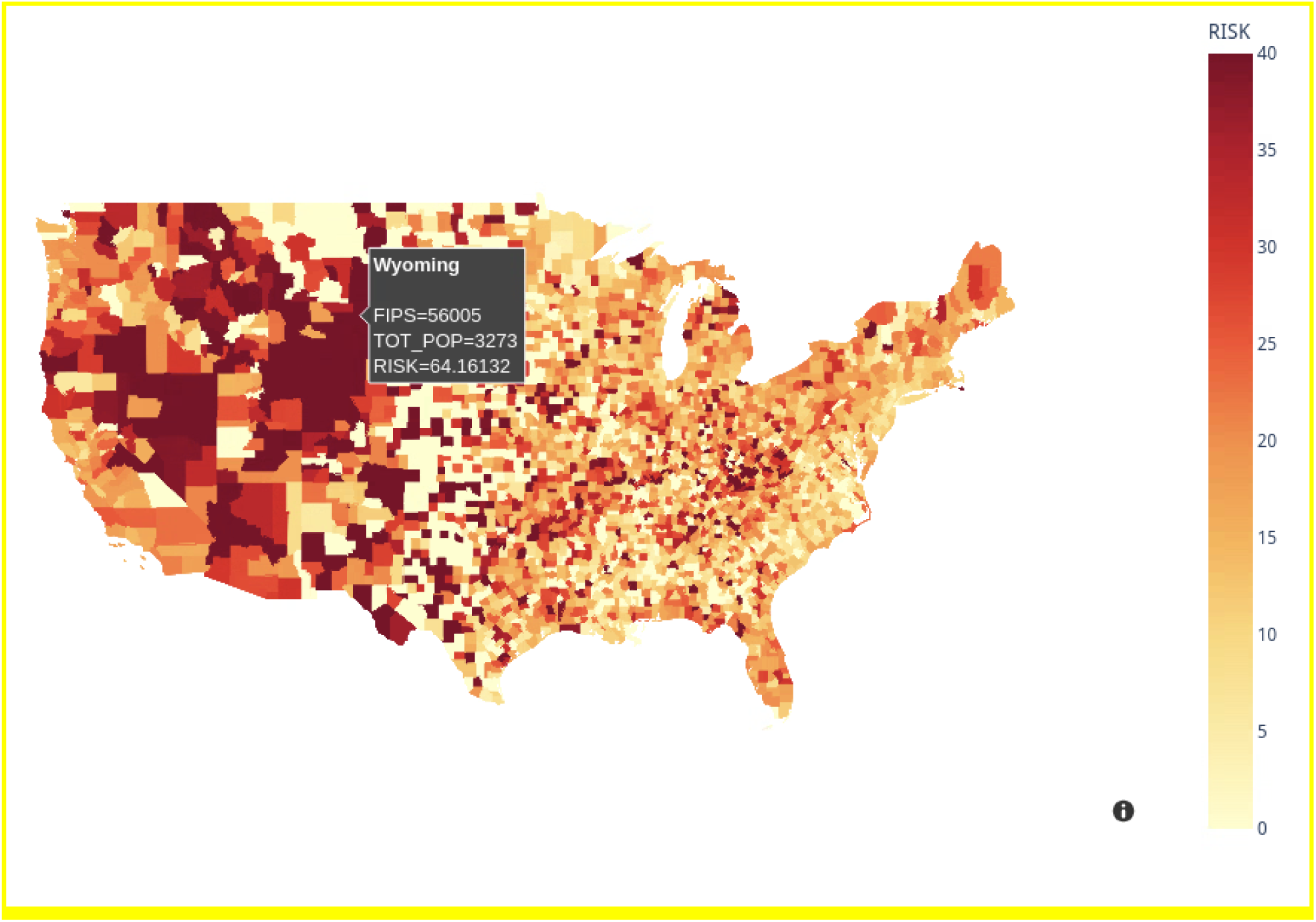

#### High Firearm Suicide Vulnerability Areas Based on Individual VHA Data

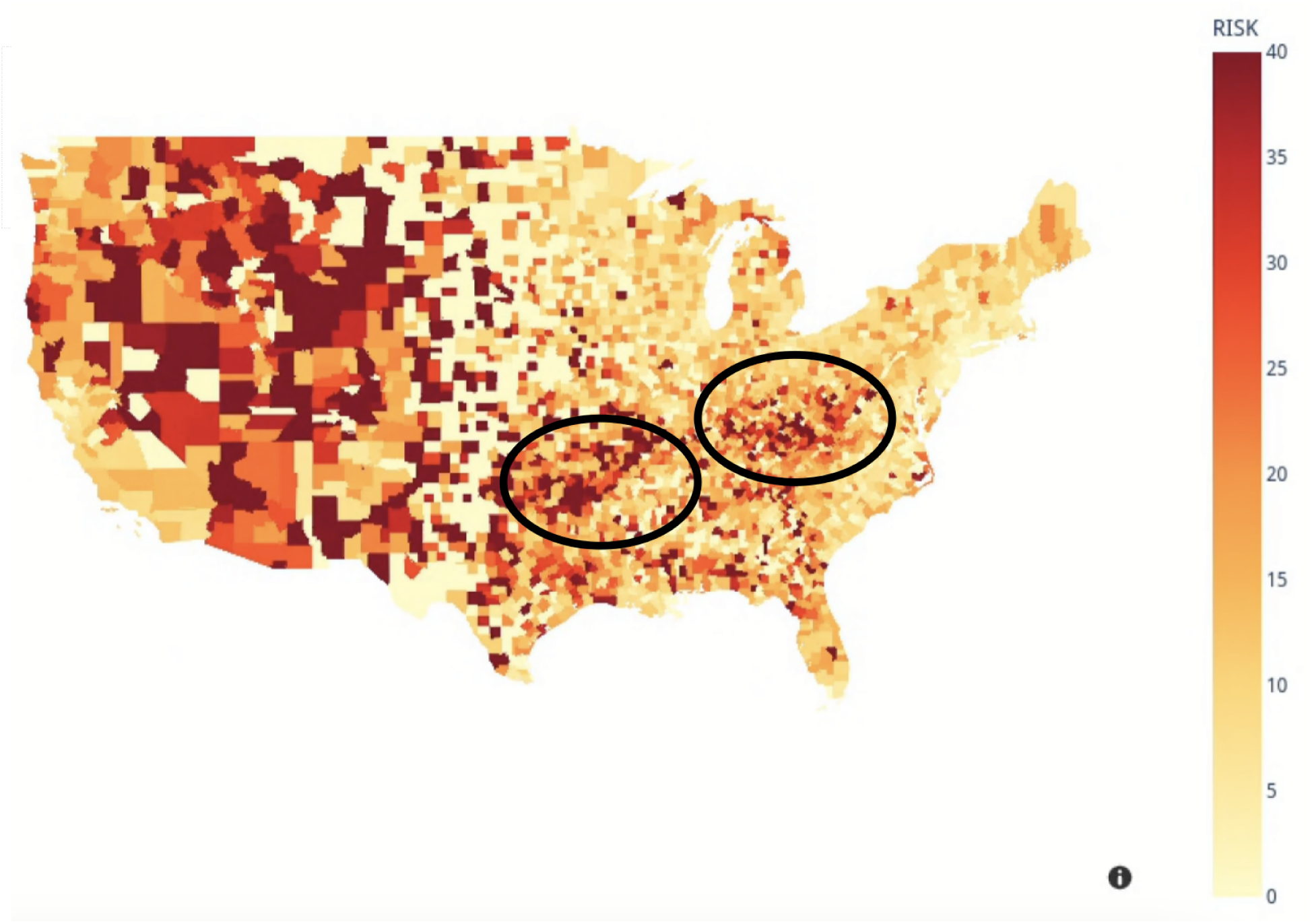

#### High Nonfirearm Suicide Vulnerability Areas Based on Individual VHA Data

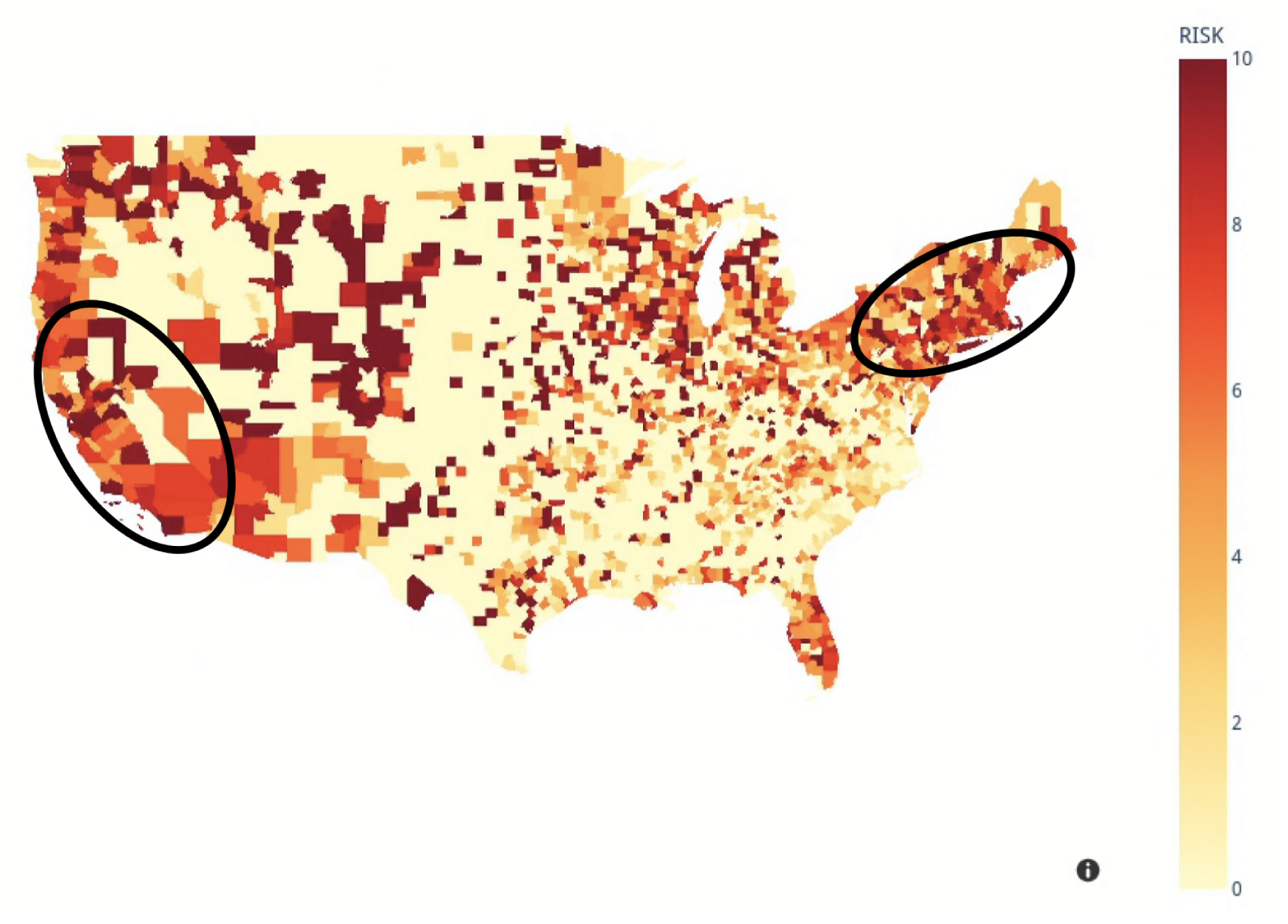

